# REACT-1 round 8 final report: high average prevalence with regional heterogeneity of trends in SARS-CoV-2 infection in the community in England during January 2021

**DOI:** 10.1101/2021.01.28.21250606

**Authors:** Steven Riley, Oliver Eales, Caroline E. Walters, Haowei Wang, Kylie E. C. Ainslie, Christina Atchison, Claudio Fronterre, Peter J. Diggle, Deborah Ashby, Christl A. Donnelly, Graham Cooke, Wendy Barclay, Helen Ward, Ara Darzi, Paul Elliott

## Abstract

In early January 2021, England entered its third national lockdown of the COVID-19 pandemic to reduce numbers of deaths and pressure on healthcare services, while rapidly rolling out vaccination to healthcare workers and those most at risk of severe disease and death. REACT-1 is a survey of SARS-CoV-2 prevalence in the community in England, based on repeated cross-sectional samples of the population. Between 6th and 22nd January 2021, out of 167,642 results, 2,282 were positive giving a weighted national prevalence of infection of 1.57% (95% CI, 1.49%, 1.66%). The R number nationally over this period was estimated at 0.98 (0.92, 1.04). Prevalence remained high throughout, but with suggestion of a decline at the end of the study period. The average national trend masked regional heterogeneity, with robustly decreasing prevalence in one region (South West) and increasing prevalence in another (East Midlands). Overall prevalence at regional level was highest in London at 2.83% (2.53%, 3.16%). Although prevalence nationally was highest in the low-risk 18 to 24 year old group at 2.44% (1.96%, 3.03%), it was also high in those over 65 years who are most at risk, at 0.93% (0.82%, 1.05%). Large household size, living in a deprived neighbourhood, and Black and Asian ethnicity were all associated with higher levels of infections compared to smaller households, less deprived neighbourhoods and other ethnicities. Healthcare and care home workers, and other key workers, were more likely to test positive compared to other workers. If sustained lower prevalence is not achieved rapidly in England, pressure on healthcare services and numbers of COVID-19 deaths will remain unacceptably high.

## Introduction

REACT-1 is a community survey of SARS-CoV-2 prevalence in England, based on repeated non-overlapping cross-sectional surveys of random samples of the population [1]. At each round, we collect self-administered throat and nose swabs and questionnaire data from between 120,000 and 180,000 people ages 5 years and above, over a period of ∼17 days, at approximately monthly intervals. Importantly, unlike the routine testing of symptomatic individuals, we include swabs irrespective of whether participants report symptoms, so as to estimate community-wide prevalence unbiased by testing availability or behaviour.

We have previously reported that prevalence of infection in early to mid January 2021 was the highest since the start of the REACT-1 programme in May 2020, towards the end of the first lockdown in England [2]. The high and rapidly increasing prevalence in the second wave appears partly to be due to the emergence of a more transmissible strain of SARS-CoV-2 that was first observed in southern England in September 2020 [3,4]. Our previous report mainly covered the period from 6th to 15th January 2021 during the first part of round 8 of the REACT-1 study, coinciding with implementation of a third national lockdown in England. Here we report results for the whole of round 8 from 6th to 22nd January 2021, and compare our findings with the positivity rate of swabs reported through routine surveillance of symptomatic individuals.

## Results and Discussion

We found 2,282 positives from 167,642 valid swabs in round 8 giving a weighted prevalence of 1.57% (95% CI, 1.49%, 1.66%) (Table 1). With a constant growth model, we found no strong evidence for either growth or decay averaged over the period between 6th to 22nd January 2021^1^ (Table 2, Figure 1). Over this period, we estimated R at 0.98 (0.92, 1.04) within the round, with weak evidence for non-constant growth: a logistic regression model with a smooth term gave ΔAIC ∼4 compared with one using only a constant term. Fitting a p-spline suggested prevalence was approximately constant or increasing slightly from 6th to 15th January, as previously reported [2], but may have then started to decline (Figure 2).

**Table 1.** Unweighted and weighted prevalence of swab-positivity across eight rounds of REACT-1.

| Round | Tested swabs | Positive swabs | Unweighted prevalence (95% CI) | Weighted prevalence (95% CI) | First sample | Last sample |
| --- | --- | --- | --- | --- | --- | --- |
| 1 | 120,620 | 159 | 0.13% (0.11%, 0.15%) | 0.16% (0.13%, 0.19%) | 1/5/2020 | 1/6/2020 |
| 2 | 159,199 | 123 | 0.077% (0.065%, 0.092%) | 0.088% (0.068%, 0.11%) | 19/6/2020 | 7/7/2020 |
| 3 | 162,821 | 54 | 0.033% (0.025%, 0.043%) | 0.040% (0.027%, 0.053%) | 24/7/2020 | 11/8/2020 |
| 4 | 154,325 | 137 | 0.089% (0.075%, 0.11%) | 0.13% (0.096%, 0.15%) | 20/8/2020 | 8/9/2020 |
| 5 | 174,949 | 824 | 0.47% (0.44%, 0.50%) | 0.60% (0.55%, 0.71%) | 18/9/2020 | 5/10/2020 |
| 6 | 160,175 | 1,732 | 1.08% (1.03%, 1.13%) | 1.30% (1.21%, 1.39%) | 16/10/2020 | 2/11/2020 |
| 6a | 85,965 | 863 | 1.00% (0.94%, 1.07%) | 1.28% (1.16%, 1.42%) | 16/10/2020 | 25/10/2020 |
| 6b | 74,210 | 869 | 1.17% (1.10%, 1.25%) | 1.32% (1.20%, 1.45%) | 26/10/2020* | 2/11/2020 |
| 7 | 168,181 | 1,299 | 0.77% (0.73%, 0.82%) | 0.94% (0.87%, 1.01%) | 13/11/2020 | 3/12/2020 |
| 7a | 105,122 | 821 | 0.78% (0.73%, 0.84%) | 0.96% (0.87%, 1.05%) | 13/11/2020 | 24/11/2020 |
| 7b | 63,059 | 478 | 0.76% (0.69%, 0.83%) | 0.91% (0.81%, 1.03%) | 25/11/2020* | 3/12/2020 |
| 8 | 167,642 | 2,282 | 1.36% (1.31%, 1.42%) | 1.57% (1.49%, 1.66%) | 06/01/2021** | 22/01/2021 |

\* Includes some samples from prior period
\*\* Includes some swab tests from 30th December onwards

**Table 2.** Estimates of national growth rates, doubling times and reproduction numbers for round 8.

| Round | Outcome | Growth rate | R* | Probability R>1 | Number of Positives |
| --- | --- | --- | --- | --- | --- |
| 8 | All positives | -0.003 ( -0.014 , 0.007 ) | 0.98 ( 0.92 , 1.04 ) | 0.27 | 2253** |
|  | Non-symptomatics | -0.012 ( -0.031 , 0.006 ) | 0.92 ( 0.81 , 1.04 ) | 0.10 | 742 |
|  | Positive for both E and N genes | -0.007 ( -0.020 , 0.006 ) | 0.95 ( 0.88 , 1.04 ) | 0.13 | 1476 |
|  | Positive for both E and N genes or positive only for N gene with CT 35 or less | -0.007 ( -0.019 , 0.004 ) | 0.95 ( 0.89 , 1.02 ) | 0.09 | 1946 |
|  | Positive for both E and N genes or positive only for N gene with CT 30 or less | -0.007 ( -0.020 , 0.006 ) | 0.96 ( 0.88 , 1.04 ) | 0.13 | 1491 |
|  | Positive for both E and N genes or positive only for N gene with CT 27 or less | -0.007 ( -0.020 , 0.006 ) | 0.96 ( 0.88 , 1.04 ) | 0.14 | 1477 |

\* Doubling/Halving time not shown because R close to 1
\*\* Discrepancy with numbers in table 1 due to positives missing a date variable not being included in the analysis

**Figure 1.**
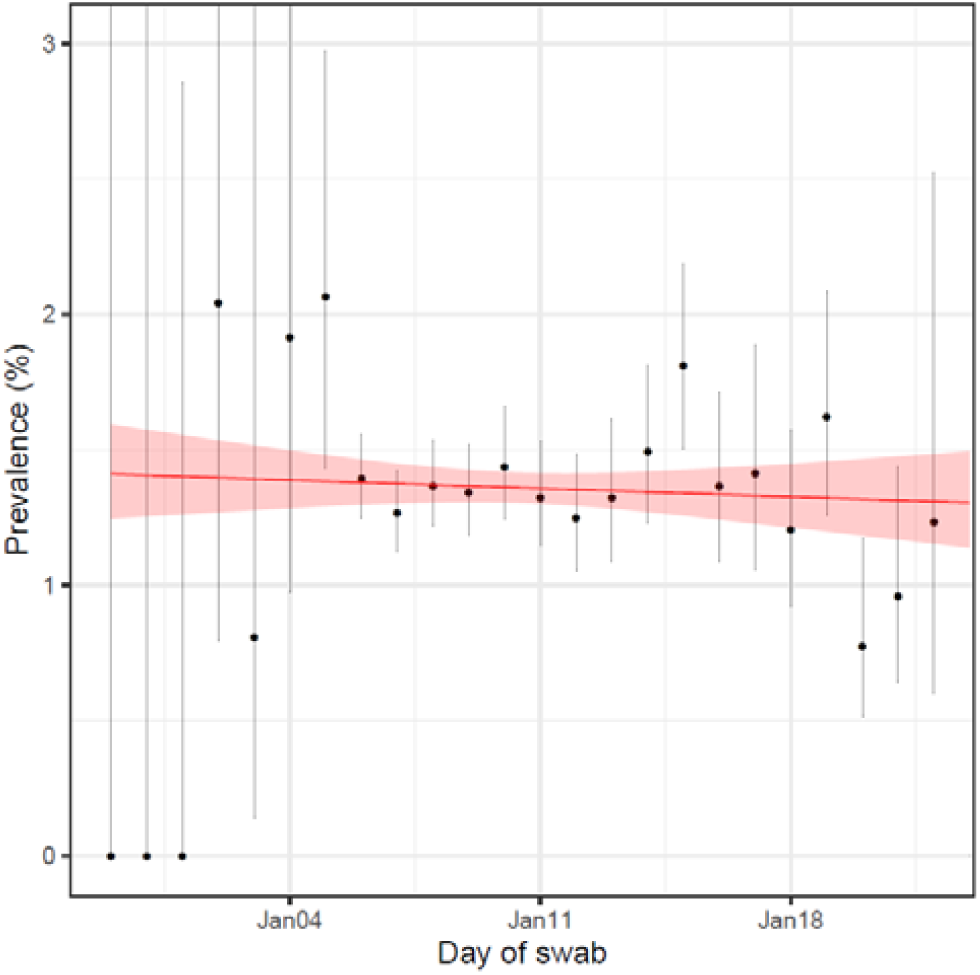
Constant growth rate model fit to REACT-1 data for England for round 8. Shaded area shows the central 95% posterior credible interval.

**Figure 2.**
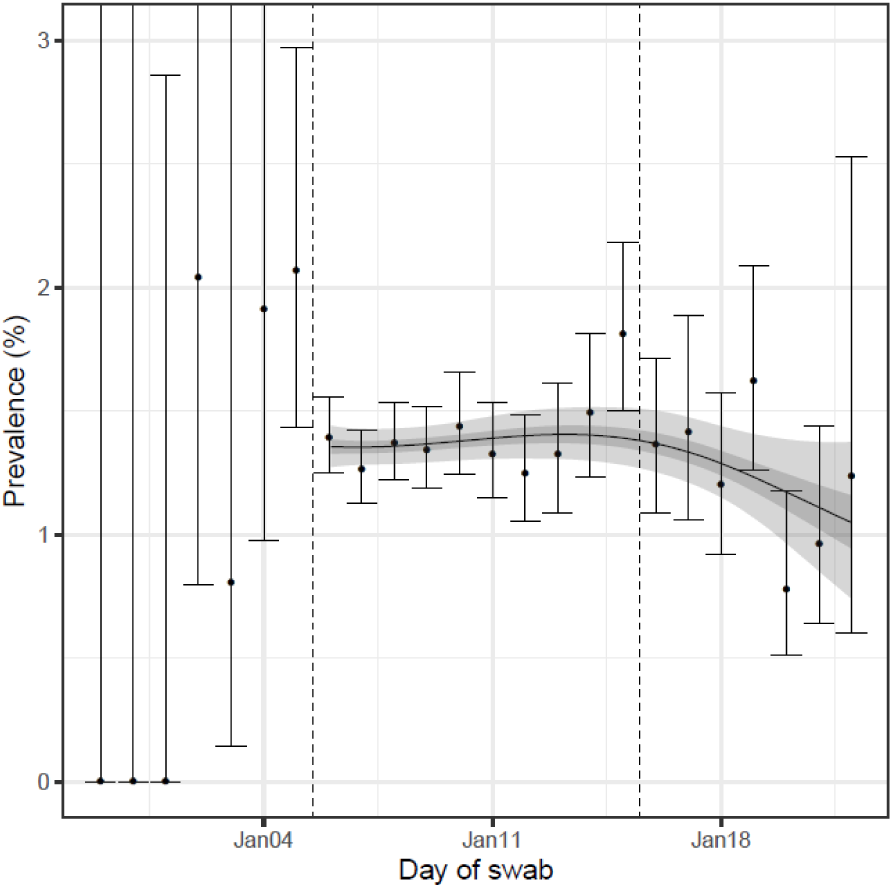
Prevalence of national swab-positivity for England estimated using a p-spline fit to all 8 rounds of data (only shown for duration of latest round) with central 50% (dark grey) and 95% (light grey) posterior credible intervals. Points show daily estimated unweighted prevalence with vertical solid lines showing 95% confidence intervals. Vertical dashed lines show the official start of round 8 and the last day of data included in the round 8a analyses.

Mobility data from the Facebook app suggest that there was a marked decrease in activity over the Christmas period and at the end of December 2020, followed by a rise in early January 2021 (Figure 3). These mobility patterns may help explain why there was an apparent fall in prevalence from a peak in late December 2020 observed by the Office for National Statistics Coronavirus Infection Survey (ONS-CIS) [5] (when REACT-1 was not collecting data). Compared with the first national lockdown, the absence of a sharp decline in prevalence during the third lockdown may be explained by a combination of: higher average levels of mobility; more people allowed to physically attend their workplace; a larger number of children eligible to attend school [6]; and a higher intrinsic transmissibility of the new variant [3,4]. We note that although there is suggestion in our data of a decline in prevalence since the 16th January, this would not be explained by patterns in the Facebook mobility data which have remained constant since early January. Rather, it may be due to the end of a temporary increase in household transmission driven by the start of lockdown, given that there is a higher prevalence of infection in people living in larger households (see below). The apparent recent decline may also reflect changing patterns of social interactions during lockdown.

**Figure 3.**
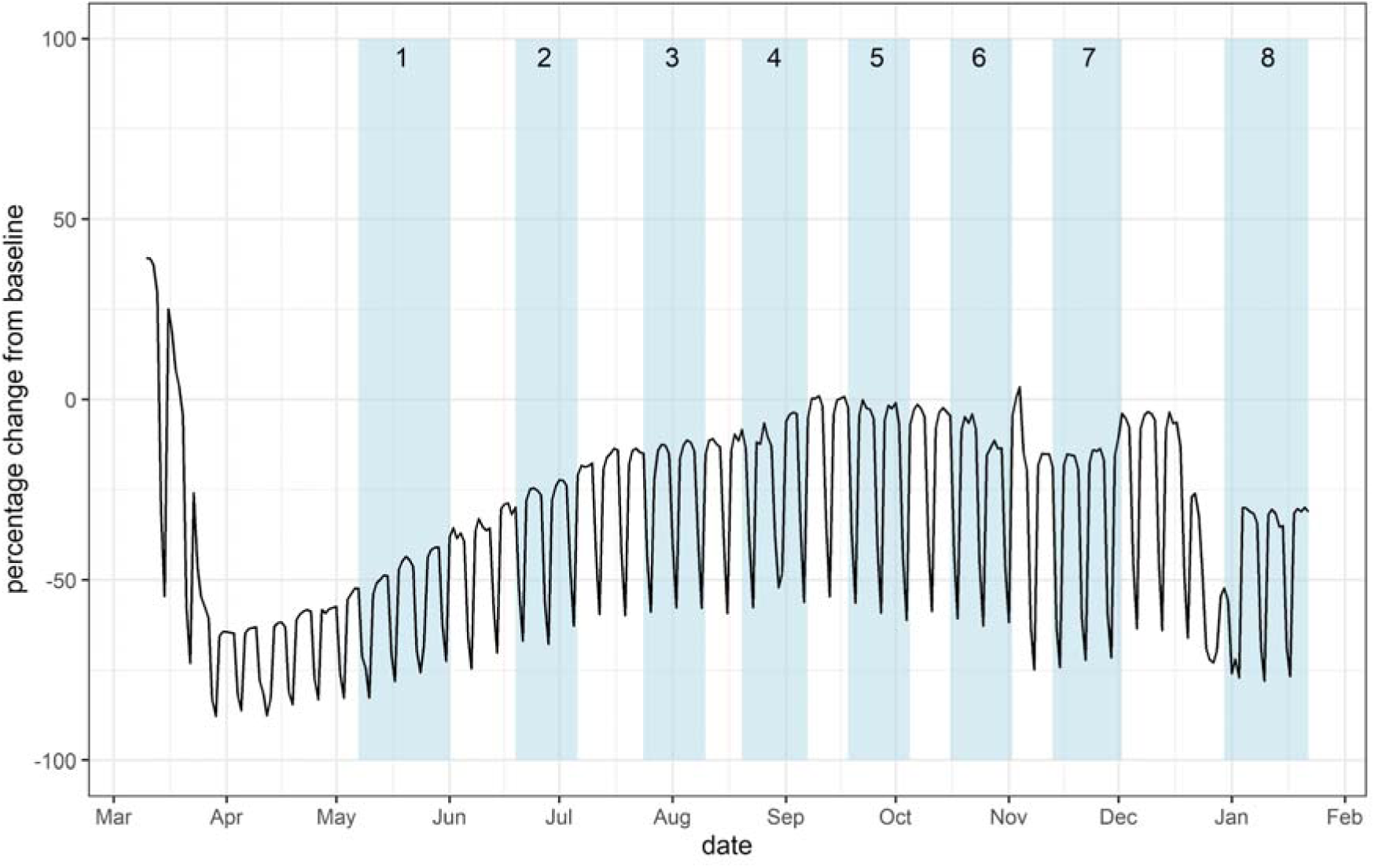
Facebook mobility data for England (black line) and the REACT-1 rounds (blue shaded regions). Baseline for mobility data is the mean movement during the week 10th - 16th March 2020 inclusive.

Regional p-splines suggest that the national average trend may mask differences in trends at the regional level (Table 3, Figure 4). Specifically, there was evidence of decline in South West, and suggestion of a decline in London and South East during the latter part of round 8. In contrast, there is evidence of an increase in prevalence in East Midlands with suggestion of increases in West Midlands and Yorkshire and The Humber. Such differences in regional trends may have contributed to higher levels of volatility in national data during previous periods of high prevalence in this pandemic [7], as they do during seasonal influenza epidemics [8]. Trends in the highest prevalence regions will have the greatest impact on the national average.

**Table 3.** Estimates of regional growth rates, doubling times and reproduction numbers for round 8.

| Round | Region | Growth rate |  | R | Probability R>1 | Halving (-) / Doubling (+) time |
| --- | --- | --- | --- | --- | --- | --- |
| 8 | East Midlands | 0.030 | ( -0.003 , 0.063 ) | 1.20 ( 0.98 , 1.45 ) | 0.96 | 22.8 ( * , 11.0 ) |
|  | West Midlands | 0.009 | ( -0.026 , 0.044 ) | 1.06 ( 0.84 , 1.30 ) | 0.70 | * ( -26.7 , 15.8 ) |
|  | East of England | -0.003 | ( -0.028 , 0.022 ) | 0.98 ( 0.83 , 1.14 ) | 0.41 | * ( -24.6 , 32.1 ) |
|  | London | -0.005 | ( -0.028 , 0.016 ) | 0.97 ( 0.83 , 1.11 ) | 0.32 | * ( -24.9 , 42.3 ) |
|  | North West | -0.009 | ( -0.045 , 0.025 ) | 0.94 ( 0.74 , 1.16 ) | 0.30 | * ( -15.4 , 28.0 ) |
|  | North East | 0.005 | ( -0.057 , 0.063 ) | 1.03 ( 0.67 , 1.45 ) | 0.56 | * ( -12.1 , 11.0 ) |
|  | South East | -0.012 | ( -0.034 , 0.009 ) | 0.92 ( 0.80 , 1.06 ) | 0.13 | * ( -20.4 , * ) |
|  | South West | -0.082 | ( -0.129 , -0.037 ) | 0.55 ( 0.36 , 0.78 ) | <0.01 | -8.5 ( -5.4 , -18.8 ) |
|  | Yorkshire ** | 0.014 | ( -0.040 , 0.063 ) | 1.09 ( 0.76 , 1.45 ) | 0.70 | 49.8 ( -17.3 , 11.0 ) |

\* Doubling/Halving time had an estimated magnitude greater than 50 days and so represented approximately constant prevalence
\*\* Yorkshire and The Humber

**Figure 4.**
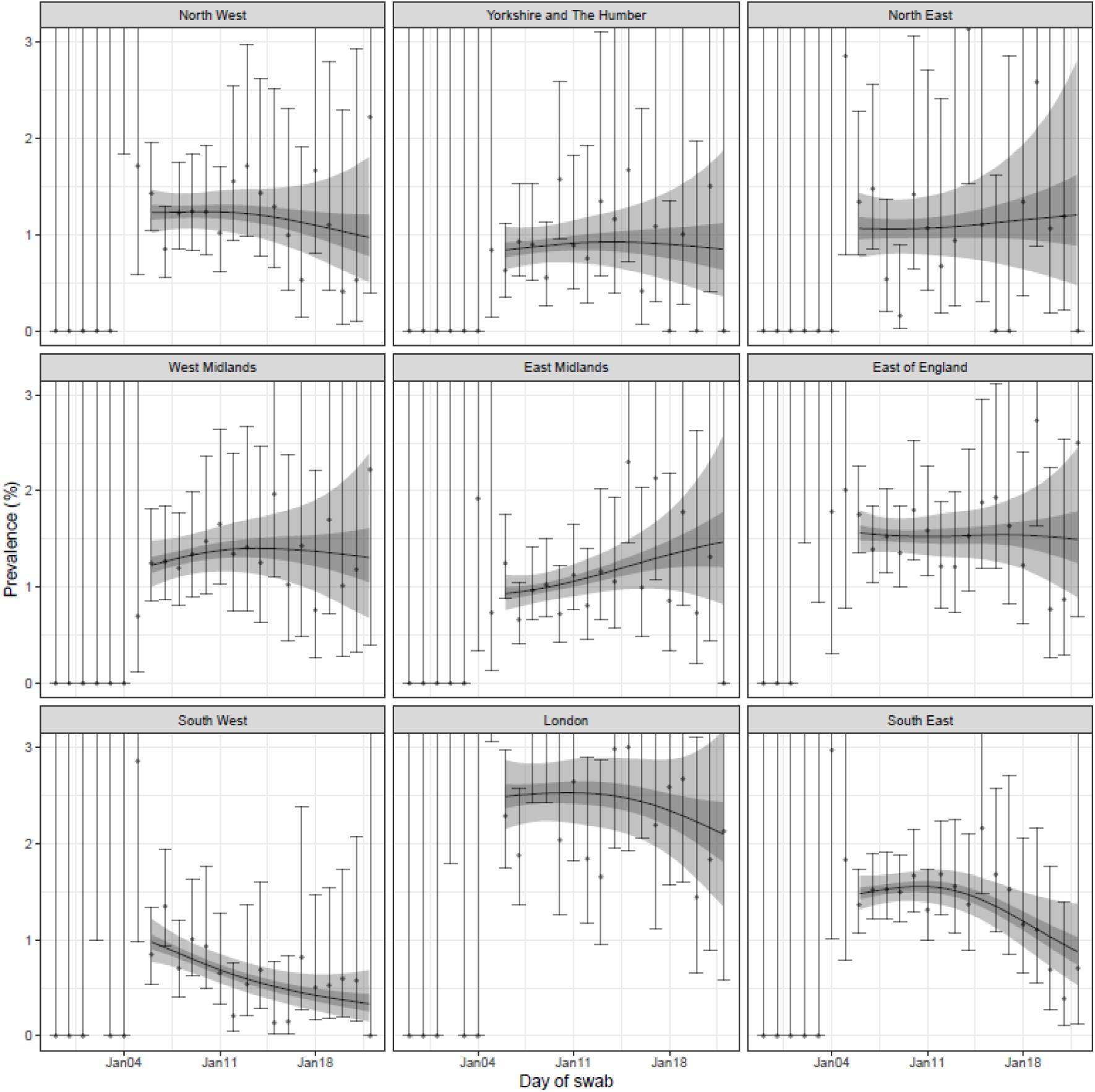
Prevalence of swab-positivity for each region of England estimated using a p-spline (with a constant second-order random walk prior distribution) fit to all 8 rounds of data (only shown for the duration of round 8) with central 50% (dark grey) and 95% (light grey) posterior credible intervals. Points show daily estimated unweighted prevalence with vertical solid lines showing 95% confidence intervals.

Regional patterns of prevalence estimated for this round of REACT-1 share key features with regional patterns of PCR-positivity from routine surveillance data [9] (Figure 5). Both data streams appear to be declining in North West, South West, London and South East. Both appear to be either level or increasing in the remaining regions. Also, in the routine surveillance data, all regions show a clear inflection point in early January which is consistent with either a plateau or substantial reduction in the rate of decline of prevalence at the time of the New Year increase in mobility (Figure 3). PCR-positivity may better reveal underlying infection patterns than case counts [9] during periods when care-seeking behaviour is changing, as has been seen in models of influenza transmission [10].

**Figure 5.**
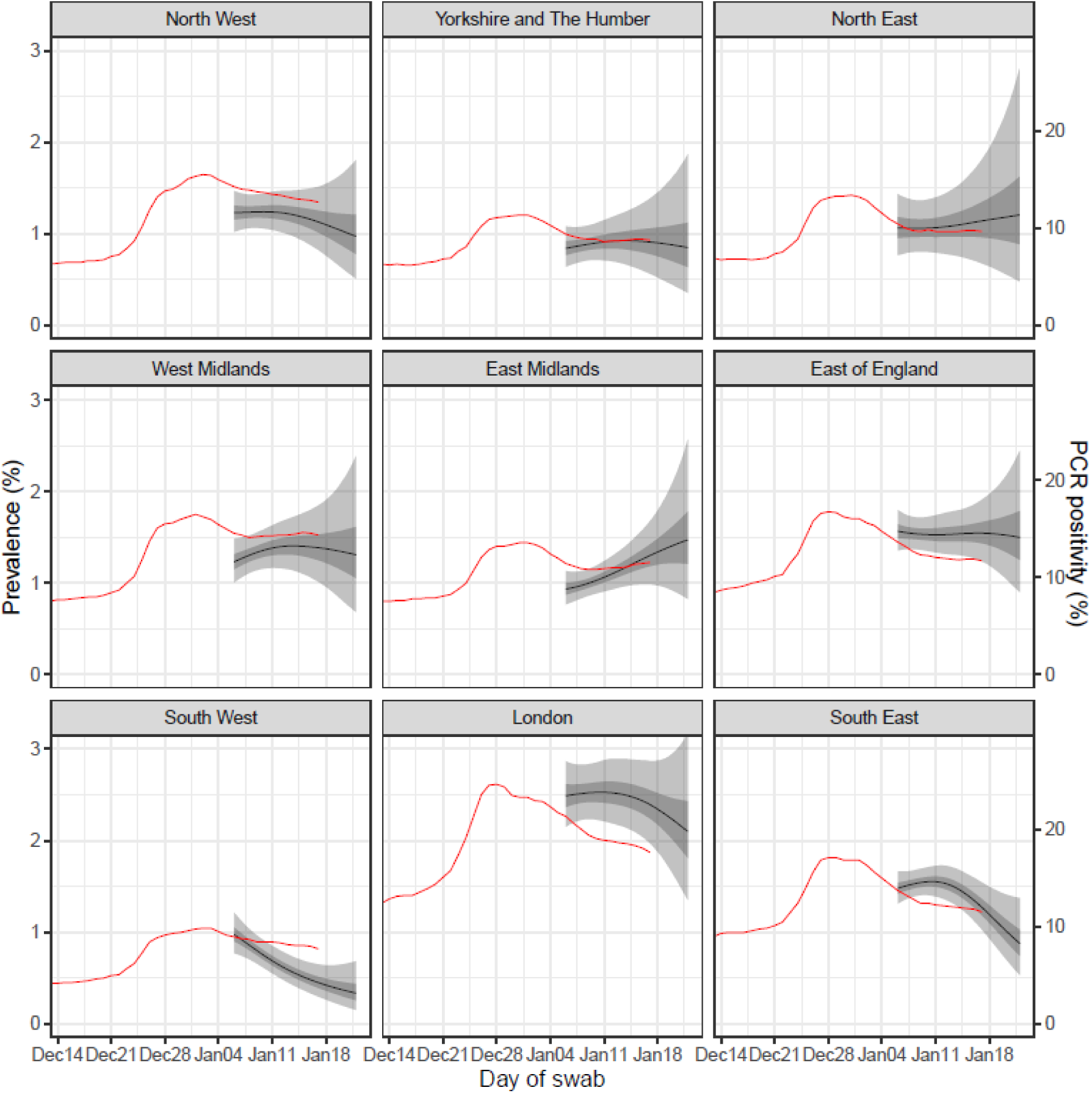
Regional comparison of REACT-1 estimated prevalence using a p-spline with central 50% (dark grey, left axis) and 95% (light grey, left axis) posterior credible intervals and routine PCR positivity for England (red, right axis) averaged over 7 days, plotted at the midpoint of the interval.

As a sensitivity analysis, we filtered the posterior distribution of prevalence trajectories, keeping only those that were consistent with the ONS-CIS estimated prevalence of 2.06% in the last 5 days of December [5] (Figure 6). This constraint did not materially affect the shape of the p-splines from January 6th, suggesting that the shape of the p-spline was robust to the absence of observations in our study during the late December 2020 peak when REACT-1 was not in the field.

**Figure 6.**
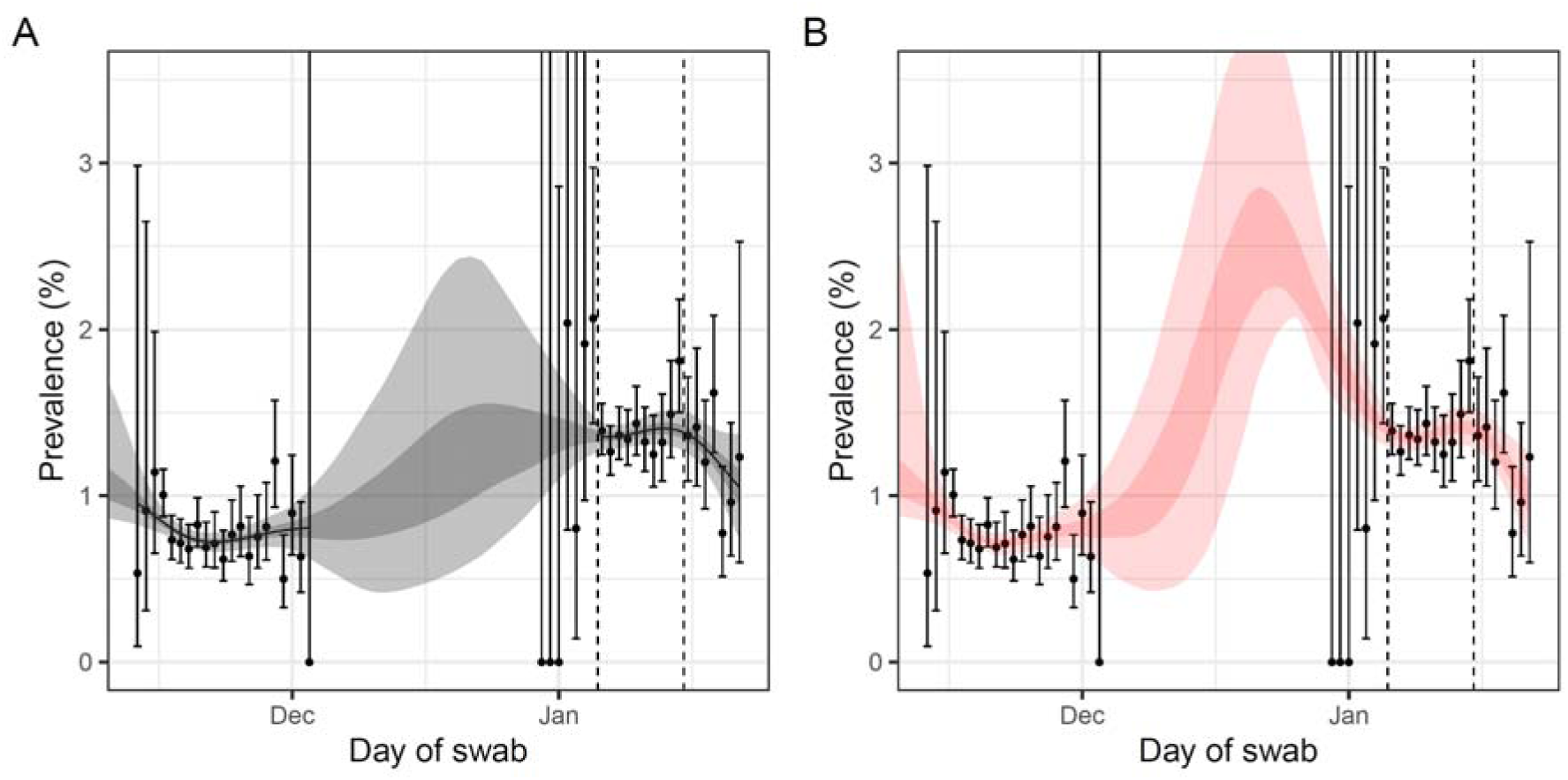
**A** Prevalence of national swab-positivity for England for rounds 7 and 8 of REACT-1 estimated using a p-spline for with central 50% (dark grey, red) and 95% (light grey, red) posterior credible intervals. **B** as A, but with posterior distribution filtered to keep only prevalence trajectories consistent with the Office for National Statistics Coronavirus Survey estimated prevalence of 2.06% in the last 5 days of December. Vertical dashed lines show the official start of round 8 and the last day of data included in the round 8a analyses.

As well as investigating trends in prevalence we undertook analyses of regional and socio-demographic variables that may be associated with increased odds of infection (Table 4, Figure 7). Regional prevalence was highest in London at 2.83% (2.53%, 3.16%), with prevalence greater than 2% in those aged 55 to 64 and those 65 years and over, and over 4% in those aged 13 to 17 and 18 to 24 years (Figure 8). This reflects the rapid increase in prevalence in London and surrounding areas in East of England and South East (Essex and Kent) that we first detected in early December 2020 [7].

**Table 4.**
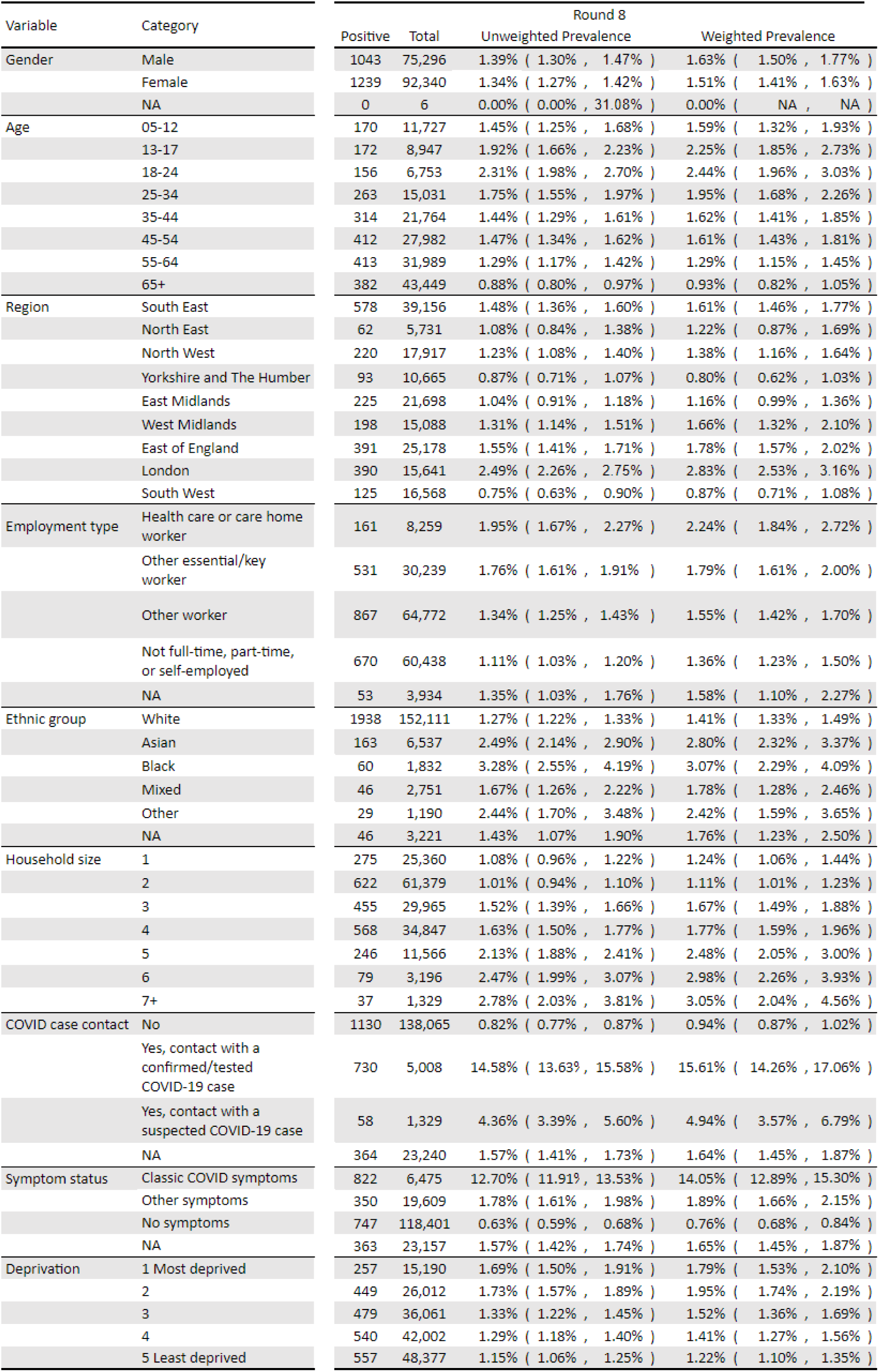
Unweighted and weighted prevalence of swab-positivity for round 8.

| Variable | Category | Round 8 |  |  |  |
| --- | --- | --- | --- | --- | --- |
|  |  | Positive | Total | Unweighted Prevalence | Weighted Prevalence |
| Gender | Male | 1043 | 75,296 | 1.39% ( 1.30% , 1.47% ) | 1.63% ( 1.50% , 1.77% ) |
|  | Female | 1239 | 92,340 | 1.34% ( 1.27% , 1.42% ) | 1.51% ( 1.41% , 1.63% ) |
|  | NA | 0 | 6 | 0.00% ( 0.00% , 31.08% ) | 0.00% ( NA , NA ) |
| Age | 05-12 | 170 | 11,727 | 1.45% ( 1.25% , 1.68% ) | 1.59% ( 1.32% , 1.93% ) |
|  | 13-17 | 172 | 8,947 | 1.92% ( 1.66% , 2.23% ) | 2.25% ( 1.85% , 2.73% ) |
|  | 18-24 | 156 | 6,753 | 2.31% ( 1.98% , 2.70% ) | 2.44% ( 1.96% , 3.03% ) |
|  | 25-34 | 263 | 15,031 | 1.75% ( 1.55% , 1.97% ) | 1.95% ( 1.68% , 2.26% ) |
|  | 35-44 | 314 | 21,764 | 1.44% ( 1.29% , 1.61% ) | 1.62% ( 1.41% , 1.85% ) |
|  | 45-54 | 412 | 27,982 | 1.47% ( 1.34% , 1.62% ) | 1.61% ( 1.43% , 1.81% ) |
|  | 55-64 | 413 | 31,989 | 1.29% ( 1.17% , 1.42% ) | 1.29% ( 1.15% , 1.45% ) |
|  | 65+ | 382 | 43,449 | 0.88% ( 0.80% , 0.97% ) | 0.93% ( 0.82% , 1.05% ) |
| Region | South East | 578 | 39,156 | 1.48% ( 1.36% , 1.60% ) | 1.61% ( 1.46% , 1.77% ) |
|  | North East | 62 | 5,731 | 1.08% ( 0.84% , 1.38% ) | 1.22% ( 0.87% , 1.69% ) |
|  | North West | 220 | 17,917 | 1.23% ( 1.08% , 1.40% ) | 1.38% ( 1.16% , 1.64% ) |
|  | Yorkshire and The Humber | 93 | 10,665 | 0.87% ( 0.71% , 1.07% ) | 0.80% ( 0.62% , 1.03% ) |
|  | East Midlands | 225 | 21,698 | 1.04% ( 0.91% , 1.18% ) | 1.16% ( 0.99% , 1.36% ) |
|  | West Midlands | 198 | 15,088 | 1.31% ( 1.14% , 1.51% ) | 1.66% ( 1.32% , 2.10% ) |
|  | East of England | 391 | 25,178 | 1.55% ( 1.41% , 1.71% ) | 1.78% ( 1.57% , 2.02% ) |
|  | London | 390 | 15,641 | 2.49% ( 2.26% , 2.75% ) | 2.83% ( 2.53% , 3.16% ) |
|  | South West | 125 | 16,568 | 0.75% ( 0.63% , 0.90% ) | 0.87% ( 0.71% , 1.08% ) |
| Employment type | Health care or care home worker | 161 | 8,259 | 1.95% ( 1.67% , 2.27% ) | 2.24% ( 1.84% , 2.72% ) |
|  | Other essential/key worker | 531 | 30,239 | 1.76% ( 1.61% , 1.91% ) | 1.79% ( 1.61% , 2.00% ) |
|  | Other worker | 867 | 64,772 | 1.34% ( 1.25% , 1.43% ) | 1.55% ( 1.42% , 1.70% ) |
|  | Not full-time, part-time, or self-employed | 670 | 60,438 | 1.11% ( 1.03% , 1.20% ) | 1.36% ( 1.23% , 1.50% ) |
|  | NA | 53 | 3,934 | 1.35% ( 1.03% , 1.76% ) | 1.58% ( 1.10% , 2.27% ) |
| Ethnic group | White | 1938 | 152,111 | 1.27% ( 1.22% , 1.33% ) | 1.41% ( 1.33% , 1.49% ) |
|  | Asian | 163 | 6,537 | 2.49% ( 2.14% , 2.90% ) | 2.80% ( 2.32% , 3.37% ) |
|  | Black | 60 | 1,832 | 3.28% ( 2.55% , 4.19% ) | 3.07% ( 2.29% , 4.09% ) |
|  | Mixed | 46 | 2,751 | 1.67% ( 1.26% , 2.22% ) | 1.78% ( 1.28% , 2.46% ) |
|  | Other | 29 | 1,190 | 2.44% ( 1.70% , 3.48% ) | 2.42% ( 1.59% , 3.65% ) |
|  | NA | 46 | 3,221 | 1.43% 1.07% 1.90% | 1.76% ( 1.23% , 2.50% ) |
| Household size | 1 | 275 | 25,360 | 1.08% ( 0.96% , 1.22% ) | 1.24% ( 1.06% , 1.44% ) |
|  | 2 | 622 | 61,379 | 1.01% ( 0.94% , 1.10% ) | 1.11% ( 1.01% , 1.23% ) |
|  | 3 | 455 | 29,965 | 1.52% ( 1.39% , 1.66% ) | 1.67% ( 1.49% , 1.88% ) |
|  | 4 | 568 | 34,847 | 1.63% ( 1.50% , 1.77% ) | 1.77% ( 1.59% , 1.96% ) |
|  | 5 | 246 | 11,566 | 2.13% ( 1.88% , 2.41% ) | 2.48% ( 2.05% , 3.00% ) |
|  | 6 | 79 | 3,196 | 2.47% ( 1.99% , 3.07% ) | 2.98% ( 2.26% , 3.93% ) |
|  | 7+ | 37 | 1,329 | 2.78% ( 2.03% , 3.81% ) | 3.05% ( 2.04% , 4.56% ) |
| COVID case contact | No | 1130 | 138,065 | 0.82% ( 0.77% , 0.87% ) | 0.94% ( 0.87% , 1.02% ) |
|  | Yes, contact with a confirmed/tested COVID-19 case | 730 | 5,008 | 14.58% ( 13.63% , 15.58% ) | 15.61% ( 14.26% , 17.06% ) |
|  | Yes, contact with a suspected COVID-19 case | 58 | 1,329 | 4.36% ( 3.39% , 5.60% ) | 4.94% ( 3.57% , 6.79% ) |
|  | NA | 364 | 23,240 | 1.57% ( 1.41% , 1.73% ) | 1.64% ( 1.45% , 1.87% ) |
| Symptom status | Classic COVID symptoms | 822 | 6,475 | 12.70% ( 11.91% , 13.53% ) | 14.05% ( 12.89% , 15.30% ) |
|  | Other symptoms | 350 | 19,609 | 1.78% ( 1.61% , 1.98% ) | 1.89% ( 1.66% , 2.15% ) |
|  | No symptoms | 747 | 118,401 | 0.63% ( 0.59% , 0.68% ) | 0.76% ( 0.68% , 0.84% ) |
|  | NA | 363 | 23,157 | 1.57% ( 1.42% , 1.74% ) | 1.65% ( 1.45% , 1.87% ) |
| Deprivation | 1 Most deprived | 257 | 15,190 | 1.69% ( 1.50% , 1.91% ) | 1.79% ( 1.53% , 2.10% ) |
|  | 2 | 449 | 26,012 | 1.73% ( 1.57% , 1.89% ) | 1.95% ( 1.74% , 2.19% ) |
|  | 3 | 479 | 36,061 | 1.33% ( 1.22% , 1.45% ) | 1.52% ( 1.36% , 1.69% ) |
|  | 4 | 540 | 42,002 | 1.29% ( 1.18% , 1.40% ) | 1.41% ( 1.27% , 1.56% ) |
|  | 5 Least deprived | 557 | 48,377 | 1.15% ( 1.06% , 1.25% ) | 1.22% ( 1.10% , 1.35% ) |

**Figure 7.**
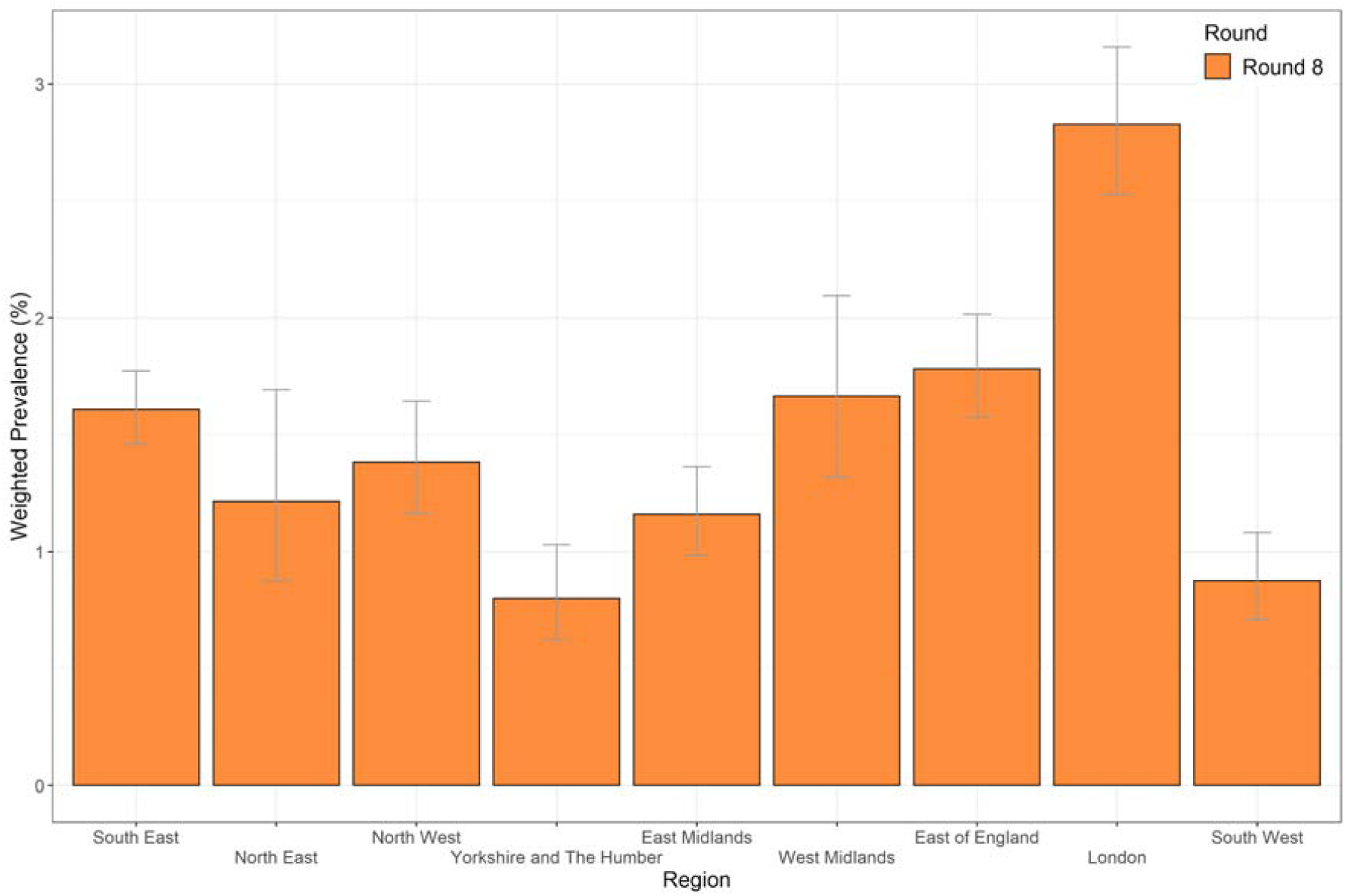
Weighted prevalence of swab-positivity by region for round 8. Bars show 95% confidence intervals.

**Figure 8.**
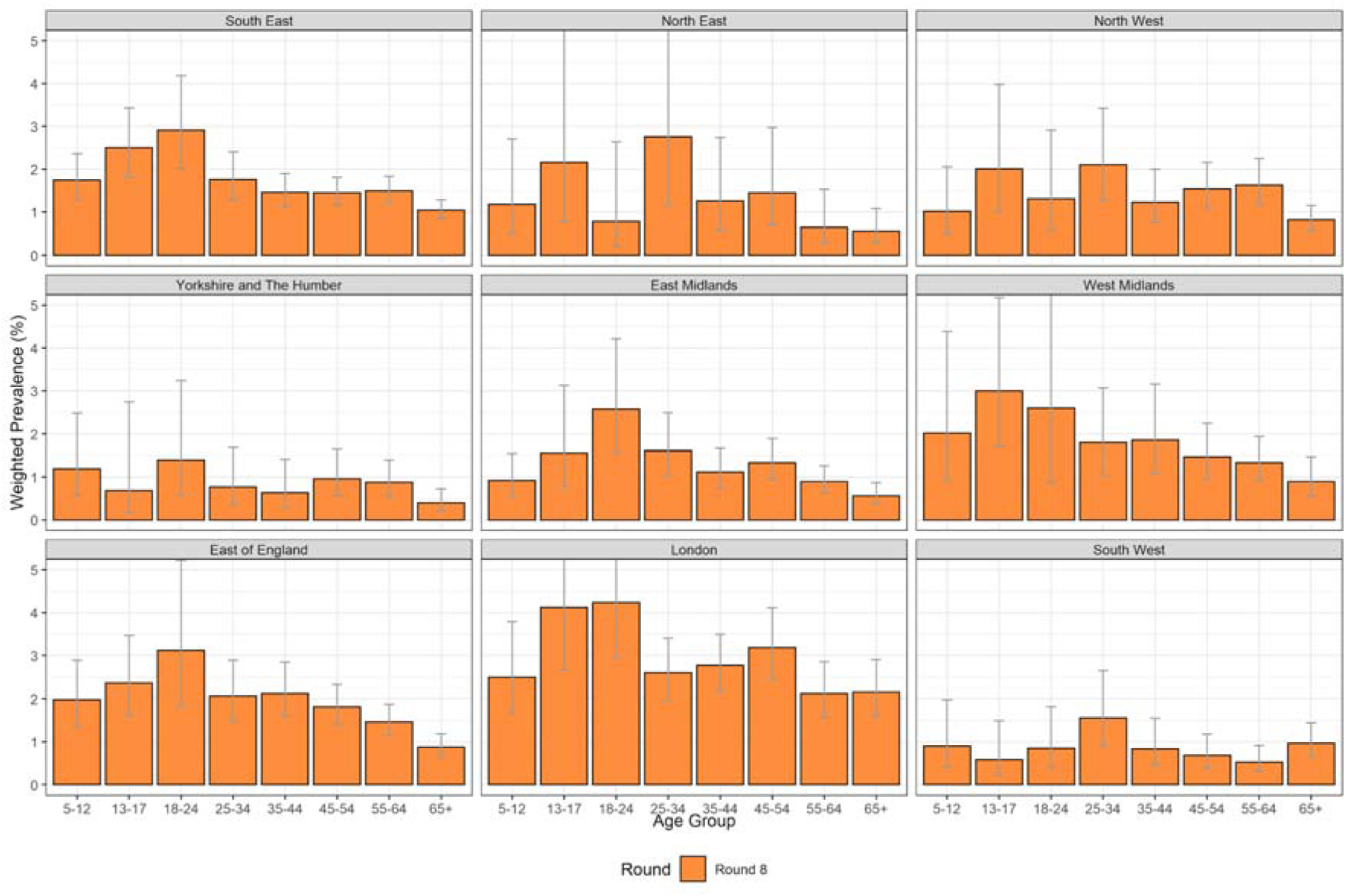
Weighted prevalence of swab-positivity by age group and region for round 8. Bars show 95% confidence intervals.

Risk of severe disease and death from SARS-CoV-2 infection is correlated strongly with increasing age [11]. Prevalence nationally was highest in 18 to 24 year olds with a weighted prevalence of 2.44% (1.96%, 3.03%), and was as high as 0.93% (0.82%, 1.05%) in those aged 65 and over (Figure 9). Previous reports have highlighted the role of transmission at younger ages [12] that has then fed through into increasing rates at older ages where people are more likely to present with severe disease.

**Figure 9.**
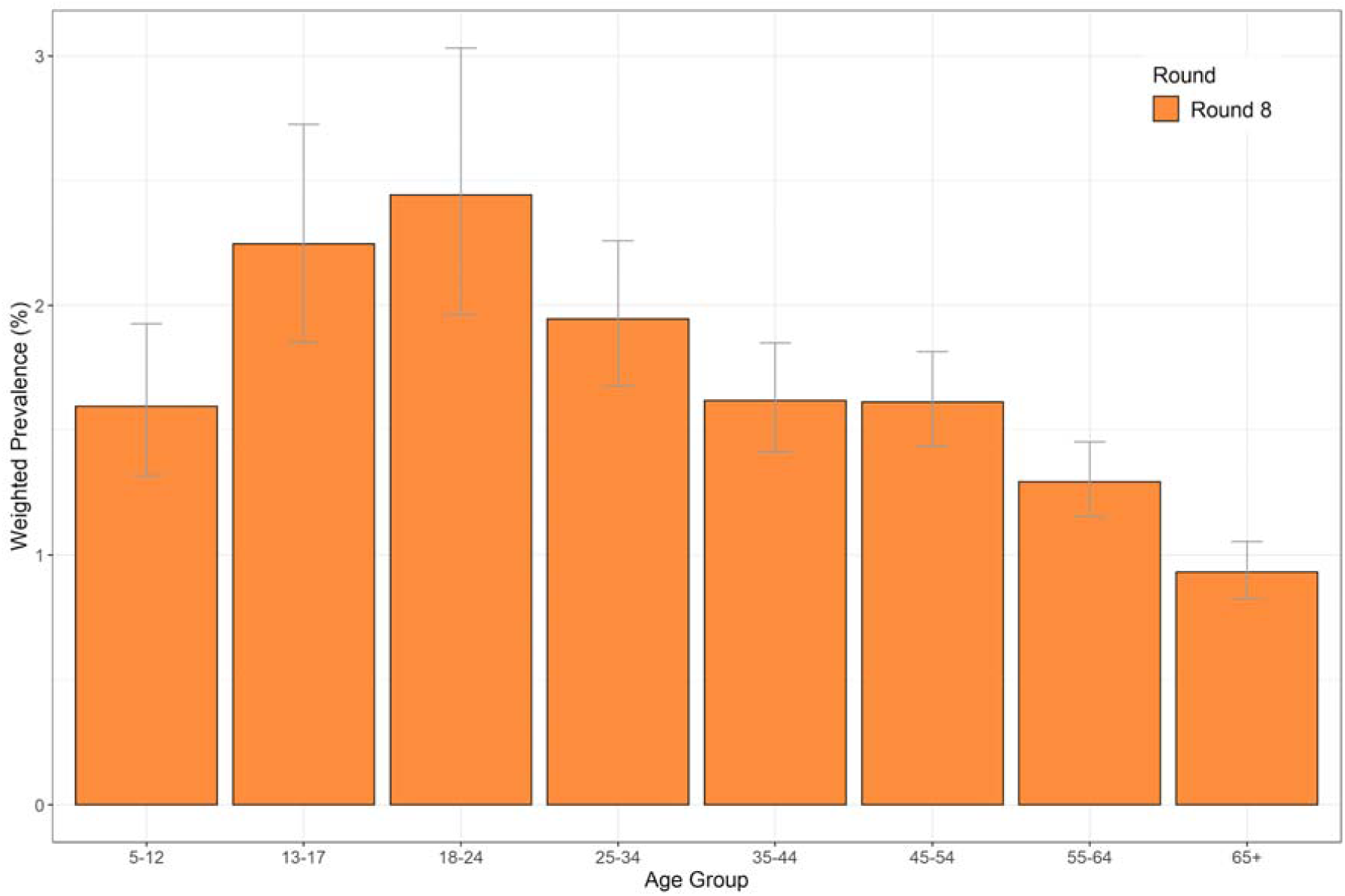
Weighted prevalence of swab-positivity by age group for round 8. Bars show 95% confidence intervals.

Large household size, living in a deprived neighbourhood, and Black and Asian ethnicity were all associated with increased prevalence (Table 4). There was a monotonic increase in prevalence from the smallest to the largest households rising from 1.24% (1.06%, 1.44%) in single-person households to 3.05% (2.04%, 4.56%) in households of seven or more people. People living in neighbourhoods in the two most deprived quintiles had prevalence of 1.79% (1.53%, 2.10%) and 1.95% (1.74%, 2.19%) compared with 1.22% (1.10%, 1.35%) for those in the least deprived. Participants of Black and Asian ethnicity had increased prevalence at 3.07% (2.29%, 4.09%) and 2.80% (2.32%, 3.37%) respectively compared with 1.41% (1.33%, 1.49%) among white participants. This is similar to data from antibody testing, indicating higher risk of infection among these minority ethnic groups [13]. Differences in prevalence for these variables were reflected in elevated odds ratios within a jointly adjusted logistic regression model (Table 5, Figure 10).

**Table 5.** Jointly adjusted odds ratios for swab-positivity by: gender, age, region, key worker status, ethnicity, household size and index of area deprivation.

| Variable | Category | Round 8 |
| --- | --- | --- |
| Gender | Male | Ref |
|  | Female | 0.95 [0.87,1.03] |
| Age Group | 5-12 | 0.94 [0.78,1.15] |
|  | 13-17 | 1.70 [1.36,2.12] |
|  | 18-24 | 1.71 [1.40,2.09] |
|  | 25-34 | 1.30 [1.10,1.54] |
|  | 35-44 | Ref |
|  | 45-54 | 1.15 [0.99,1.34] |
|  | 55-64 | 1.19 [1.02,1.39] |
|  | 65+ | 1.02 [0.85,1.22] |
| Region | North East | 0.70 [0.54,0.92] |
|  | North West | 0.78 [0.66,0.92] |
|  | Yorkshire | 0.60 [0.48,0.75] |
|  | East Midlands | 0.69 [0.59,0.81] |
|  | West Midlands | 0.86 [0.73,1.02] |
|  | East of England | 1.01 [0.89,1.16] |
|  | London | 1.47 [1.27,1.69] |
|  | South East | Ref |
|  | South West | 0.51 [0.42,0.62] |
| Key Worker Status | HCW/CHW | 1.48 [1.25,1.77] |
|  | Key worker (other) | 1.35 [1.20,1.51] |
|  | Other worker | Ref |
|  | Not FT, PT, SE | 0.90 [0.80,1.02] |
| Ethnicity | Asian | 1.35 [1.14,1.61] |
|  | Black | 1.63 [1.24,2.14] |
|  | Mixed | 1.03 [0.77,1.39] |
|  | Other | 1.39 [0.96,2.03] |
|  | White | Ref |
| Household Size | 1-2 People | Ref |
|  | 3-5 People | 1.37 [1.24,1.52] |
|  | 6+ People | 2.02 [1.64,2.50] |
| Deprivation Index Quintile | 1 - Most Deprived | Ref |
|  | 2 | 1.03 [0.88,1.21] |
|  | 3 | 0.82 [0.70,0.97] |
|  | 4 | 0.82 [0.70,0.96] |
|  | 5 - Least Deprived | 0.72 [0.61,0.84] |

**Figure 10.**
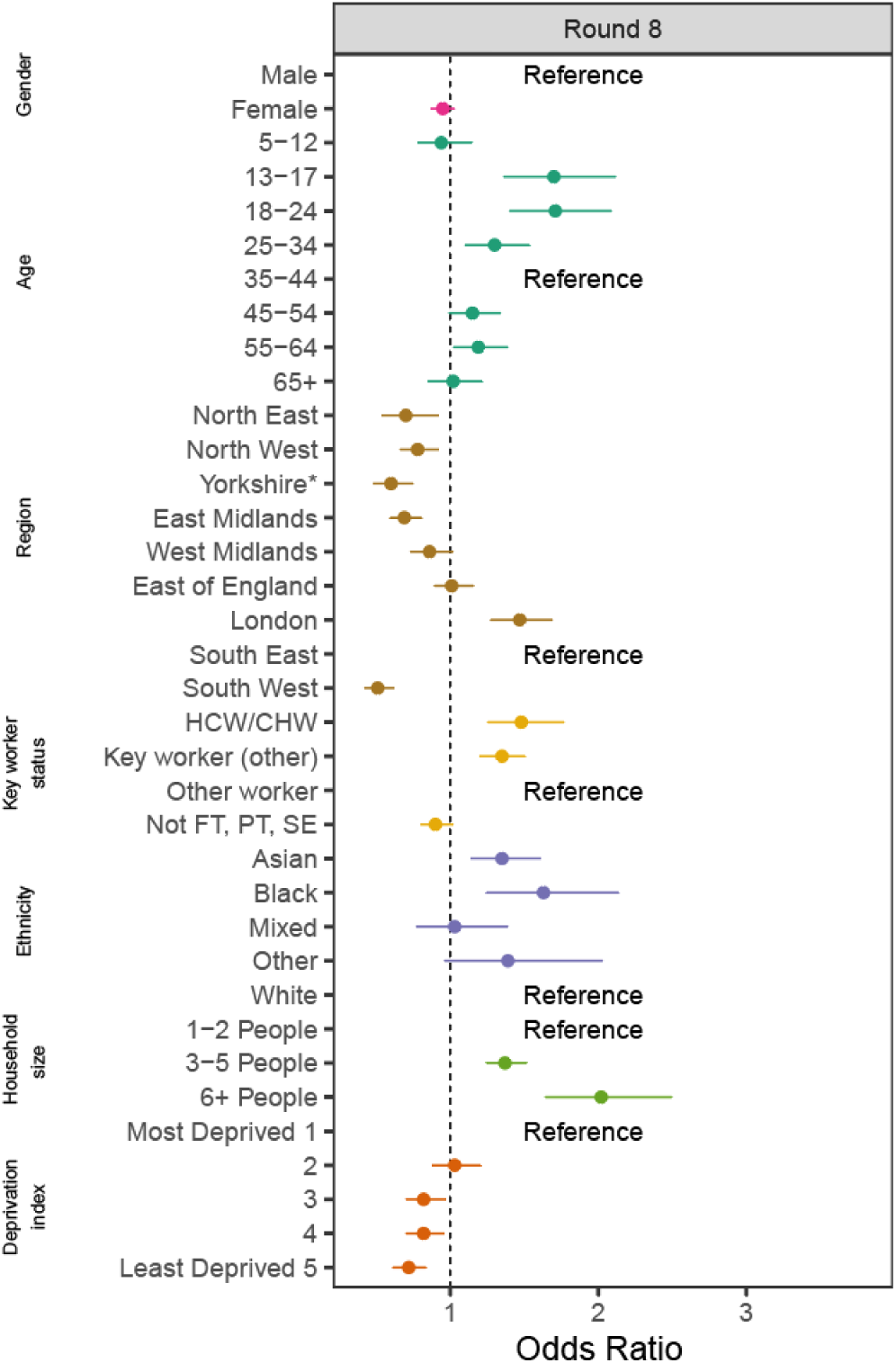
Estimated odds ratios and 95% confidence intervals for mutually-adjusted logistic regression model of swab-positivity for round 8. Models were adjusted for gender, age group, region, key worker status, ethnicity, household size, and deprivation index. The deprivation index is based on the Index of Multiple Deprivation (2019) at lower super output area. Here we group scores into quintiles, where 1 = most deprived and 5 = least deprived. HCW/CHW = healthcare or care home workers; Not FT, PT, SE = Not full-time, part-time, or self-employed. *Yorkshire and The Humber.

Both healthcare and care home workers, and other key workers, had increased odds of swab-positivity compared to other workers at 1.48 (1.25, 1.77) and 1.35 (1.20, 1.51) respectively (Table 5, Figure 10). Health care and care home workers had over five-fold odds of infection compared with other workers at the end of the first wave [14], indicating the importance of nosocomial infections in driving the epidemic at that time. This category and other key workers continue to have higher prevalence of infection than other workers which likely reflects higher occupational exposure to the virus.

We used a nearest-neighbours statistic to generate a smoothed prevalence map for round 8 at the level of lower-tier local authority (LTLA) (Figure 11). We observed marked heterogeneity with highest prevalence in London and a contiguous area radiating out into East of England and South East. There were also pockets of high prevalence in West Midlands and other regions.

**Figure 11.**
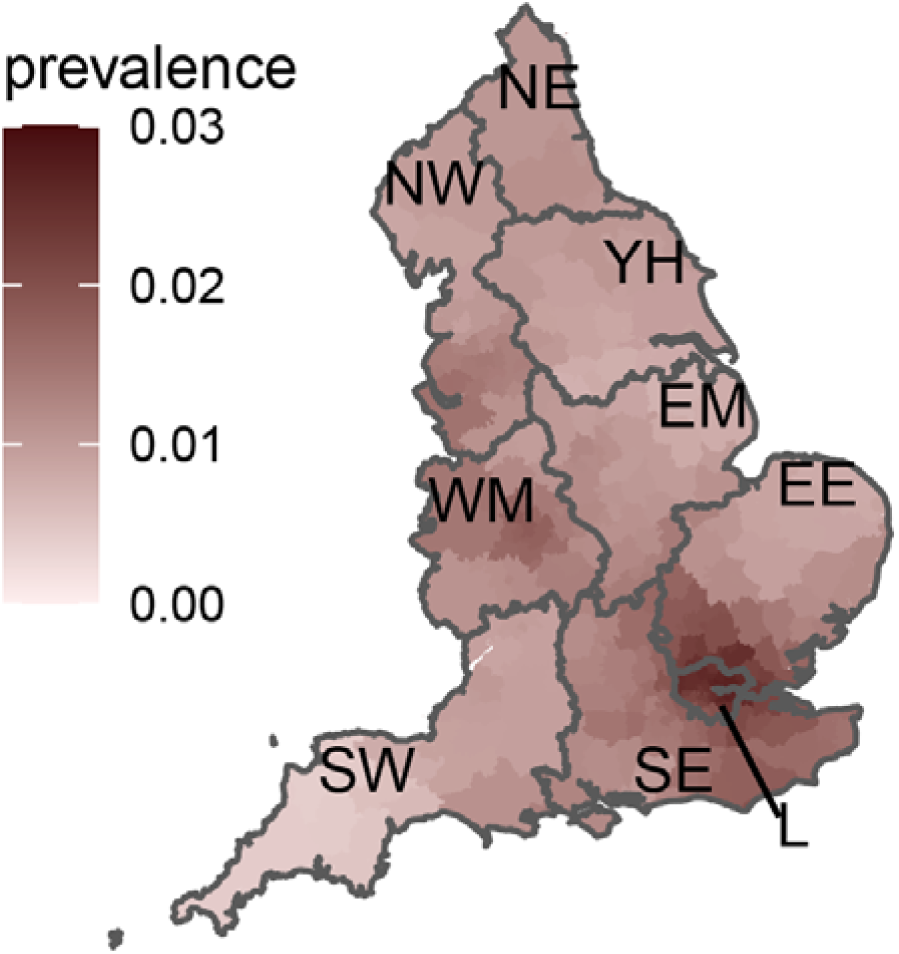
Neighbourhood prevalence of swab-positivity for round 8. Neighbourhood prevalence calculated from nearest neighbours (the median number of neighbours within 30km in the study). Average neighbourhood prevalence displayed for individual lower-tier local authorities. Regions: NE = North East, NW = North West, YH = Yorkshire and The Humber, EM = East Midlands, WM = West Midlands, EE = East of England, L = London, SE = South East, SW = South West. Data for unweighted point estimate of prevalence available in the supplementary data file.

## Conclusions

**I**n this large study of SARS-CoV-2 prevalence in the community in England, we show that prevalence in January 2021 nationally was at extremely high levels. This is being reflected in high levels of hospital admissions, intensive care admission and deaths [9]. While there was indication of a possible decline in prevalence toward the end of our study period (up to 22nd January), the levels of infection remain much higher than those seen during lockdown in May 2020 [14], with a shallower downward trajectory. Unless the prevalence of infection in the community can be lowered substantially, the extreme pressure on health services will continue over the coming weeks, and possibly months, until the vaccination programme protects sufficient numbers of at-risk individuals. In the meantime, it is essential that we continue to observe public health measures to contain the virus including social distancing, face covers, hand-washing and isolation of cases.

## Methods

REACT-1 methods are published [1]. At each round, we collect questionnaire data and self-administered throat and nose swabs (using dry swabs) which are sent at 4^°^ to 8^°^C to a single laboratory for RT-PCR. We obtain the sample from the list of National Health Service patients, selected randomly and stratified at LTLA level (n=315) to give approximately equal numbers for each LTLA.

In round 8, we sent out 761,007 registration letters to named individuals, of whom 213,746 (28.1%) registered (parent/guardian for children ages 5 to 12 years); swabs from 168,988 (79.1%) people were returned of which 1,346 (0.8%) had an invalid RT-PCR result. That left 167,642 returned swabs with a valid result, giving an overall response rate (valid swabs divided by total number of people invited) of 22.0%.

We obtain unweighted prevalence estimates from the counts of swab-positivity (based on RT-PCR) compared with the number of swabs returned; we also calculate prevalence estimates weighted to be representative of the population of England as a whole, based on age, sex, region and ethnicity. We estimate prevalence by socio-demographic and other characteristics and use multivariable logistic regression to obtain jointly adjusted estimates of the odds of swab-positivity.

Per round, we use exponential growth models to estimate time trends and reproduction number R, nationally and regionally, and obtain smoothed within-round prevalence estimates using a p-spline function with knots at 5-day intervals [15]. We evaluate geographic variation in prevalence by LTLA using a neighbourhood spatial smoothing method based on nearest neighbour up to 30 km as previously described [16].

We carry out a range of sensitivity analyses including estimation of R for different cut-points for CT values that determine swab-positivity and for non-symptomatic individuals (not reporting symptoms on the day of swab or week prior). We also compare data from the national routine programme of symptomatic testing [9] with our own data at regional level.

Statistical analyses are carried out in R [17]. We obtained research ethics approval from the South Central-Berkshire B Research Ethics Committee (IRAS ID: 283787).

## Data Availability

Supporting data for tables and figures are available either: in this Google spreadsheet https://docs.google.com/spreadsheets/d/1GouQlf0aBmzZSVG-EGMwvIuwRx-O2PDHIZ-KVlW5KLs/edit?usp=sharing; or in the inst/extdata directory of this GitHub R package https://github.com/mrc-ide/reactidd.

https://docs.google.com/spreadsheets/d/1GouQlf0aBmzZSVG-EGMwvIuwRx-O2PDHIZ-KVlW5KLs/edit?usp=sharing

## Data availability

Supporting data for tables and figures are available either: in this Google <u>spreadsheet</u>; or in the inst/extdata directory of this <u>GitHub R package</u>.

## Acknowledgements

SR, CAD acknowledge support: MRC Centre for Global Infectious Disease Analysis, National Institute for Health Research (NIHR) Health Protection Research Unit (HPRU), Wellcome Trust (200861/Z/16/Z, 200187/Z/15/Z), and Centres for Disease Control and Prevention (US, U01CK0005-01-02). GC is supported by an NIHR Professorship. PE is Director of the MRC Centre for Environment and Health (MR/L01341X/1, MR/S019669/1). PE acknowledges support from Health Data Research UK (HDR UK); the NIHR Imperial Biomedical Research Centre; NIHR HPRUs in Chemical and Radiation Threats and Hazards, and Environmental Exposures and Health; the British Heart Foundation Centre for Research Excellence at Imperial College London (RE/18/4/34215); and the UK Dementia Research Institute at Imperial (MC_PC_17114). We thank The Huo Family Foundation for their support of our work on COVID-19.

We thank key collaborators on this work – Ipsos MORI: Kelly Beaver, Sam Clemens, Gary Welch, Nicholas Gilby, Kelly Ward and Kevin Pickering; Institute of Global Health Innovation at Imperial College: Gianluca Fontana, Dr Hutan Ashrafian, Sutha Satkunarajah, Didi Thompson and Lenny Naar; Molecular Diagnostic Unit, Imperial College London: Prof. Graham Taylor; North West London Pathology and Public Health England for help in calibration of the laboratory analyses; NHS Digital for access to the NHS register; and the Department of Health and Social Care for logistic support. SR acknowledges helpful discussion with attendees of meetings of the UK Government Office for Science (GO-Science) Scientific Pandemic Influenza – Modelling (SPI-M) committee.

## Footnotes

1 includes 2,229 swabs collected from 30th December 2020 to 5th January 2021

